# Cytomegalovirus seropositivity relates inversely to cancer incidences across races and ethnicities: implications for oncoprevention

**DOI:** 10.1101/2023.08.26.23294534

**Authors:** Marko Janković, Ognjen Milićević, Milena Todorović-Balint, Irena Đunić, Biljana Mihaljević, Tanja Jovanović, Aleksandra Knežević

**Affiliations:** Institute of Microbiology and Immunology, Department of Virology, Faculty of Medicine, University of Belgrade, 1 Dr Subotića street, 11000 Belgrade, Republic of Serbia; Institute of Medical Statistics and Informatics, Faculty of Medicine, University of Belgrade, 15 dr Subotića street, 11000 Belgrade, Republic of Serbia; Clinic of Hematology, Faculty of Medicine, University Clinical Centre of Serbia, University of Belgrade, 2 dr Koste Todorovića street, 11000 Belgrade, Republic of Serbia

**Keywords:** Cancer, cytomegalovirus, oncoprotection, global, United States

## Abstract

**Background:** Race and ethnic disparities in cancer incidence rates and the prevalence of *cytomegalovirus* (CMV) are known to exist in the United States (U.S.) but also across broad geographic expanses. CMV prevalence seems to inversely contrast tumor incidence rates both in ethnic groups and globally. Is there a biological link between cancer and CMV infection? Most recent clinical results seem to certify it.

**Methods:** Global cancer data were retrieved from the World Health Organization (WHO) database. Incidence of cancer and CMV seroprevalence (73 countries) were subjected to Spearman’s correlation test. The Bayesian framework was adopted for CMV seropositivity variables. Relevant data for the U.S. were extracted from publications based on the Surveillance, Epidemiology, and End Results (SEER) registries and the National Health and Nutrition Examination Surveys (NHANES), 1988-2004.

**Results:** An inversely directed coupling between cancer and CMV seropositivity across diverse ecologies and cultural domains suggest a global oncoprotective effect of the CMV (Spearman’s *ρ* = -0.732; *p*<0.001). Rates of all cancers combined and CMV seropositivity show an opposite association (*p*<0.001) among the races and foremost U.S. ethnic groups.

**Conclusion:** The racial/ethnic incidence of cancers and CMV seropositivity are inversely proportional both in the U.S. and globally. This would support a view that CMV is a potential driver against tumorigenesis. An absence of CMV infection abrogates protection against malignant clones afforded to an infected host. Abating CMV seroprevalence may relate causally to the buildup of malignancies in U.S. and the West world countries with thriving hygiene and healthcare systems.

**Importance:** Increasing evidence substantiates the potential of cytomegalovirus (CMV) to counteract tumors, particularly in the field of anti-cancer vaccinology, leading to extended periods of survival. This research unveils a robust and inverse correlation between the prevalence of CMV and the occurrence of cancer both within the United States and on a global scale (73 countries), hinting at the ability of CMV to inhibit tumor development. Furthermore, this phenomenon remains consistent across various racial and ethnic groups within the United States and applies to a diverse range of cancer types. A notable pattern emerges: the higher the prevalence of the viral infection, the lower the incidence of tumors within a given country. These findings support existing insights from clinical and experimental investigations, underscoring the notion that this effect becomes apparent at the level of entire nations and populations worldwide.

## Introduction

Human cytomegalovirus (CMV) is a ubiquitous β-herpesvirus typically causing an asymptomatic to mild infection in healthy children and adults. In the immunocompromised patient and the unprotected fetus CMV can present as a significant cause of morbidity and mortality. After primary contact, the virus persists within the host as a lifelong infection awakening at times from dormancy and leading to reactivation and virion shedding. Over millennia of evolutionary time, CMV had interlocked in a complex interplay with the immune system of its human host, with growing evidence pointing to T lymphocytes as being galvanized by CMV and directed against specific cancer cells. Recently, we drew attention to an inversely correlated incidence of human B lymphocyte malignancies and CMV seroprevalence in a single-center clinical setting and in humans across races/ethnicities the world over (Spearman’s coefficient *ρ* = −0.625, *p* < 0.001) [13]. This hinted at a possible oncopreventive faculty of the virus (**Fig. 1**). Similar findings were described in animal *in vitro* tissue models [6-8] covering several oncological diseases [9-11,12]. This indicates that CMV infection may bestow a capacity on its host to resist carcinogenesis somewhat more effectively than does a normal immunity in an uninfected subject. In effect, CMV and cancer may be profoundly connected. Virus/host interaction has not been investigated in a racial/ethnic background or across the globe. Inquiring into this association we used the wealth of data derived from authorized literature based on the U.S. registries and global publications (World Health Association [WHO]).

**Figure 1.**
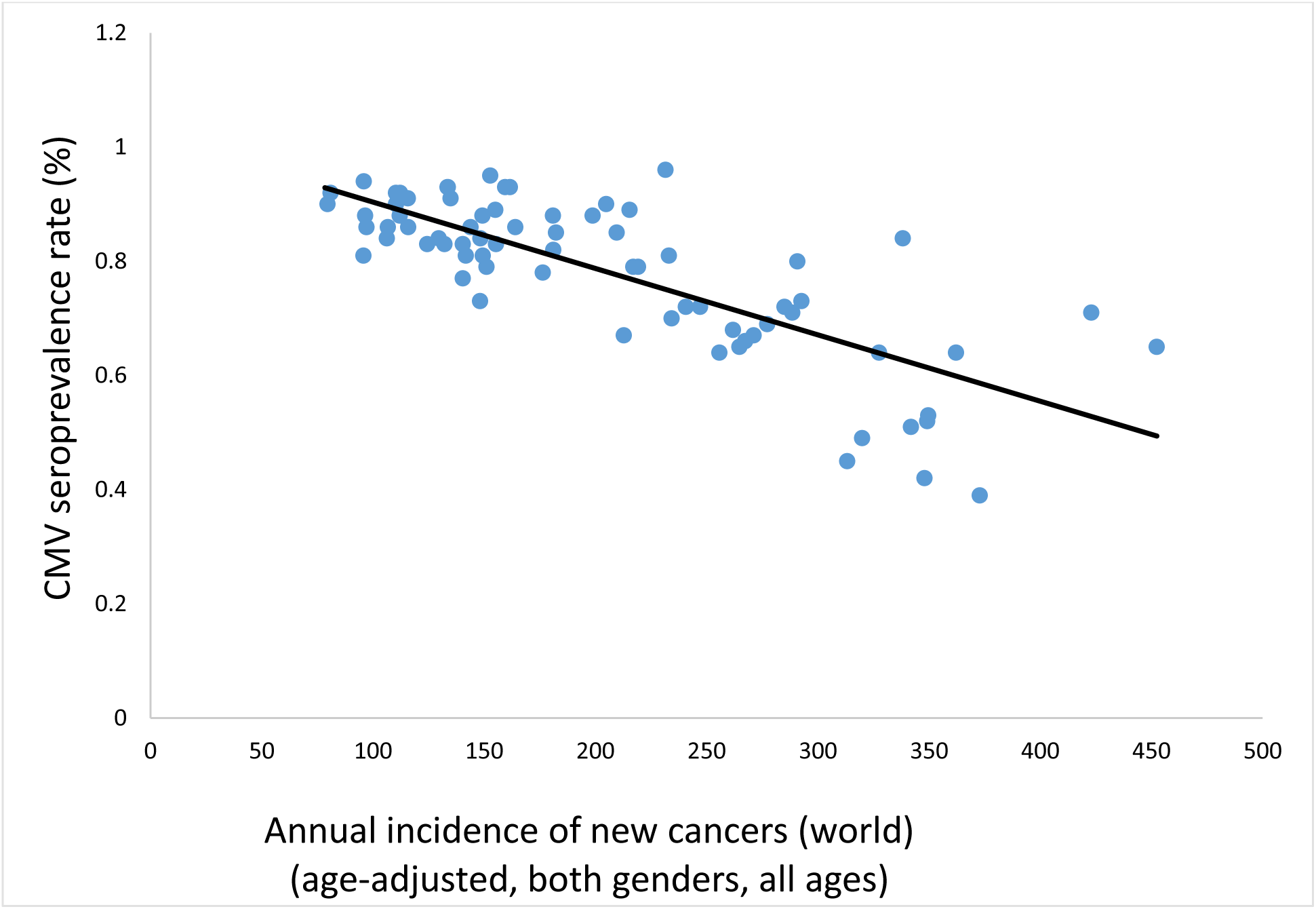
New cancer (all invasive types combined, all sites)/10^5^ country specific population plotted against CMV seropositivity rates [28]. Inverse relationship over 73 countries [51] (Spearman’s *ρ* = -0.732; *p*<0.001), signals a globally pervasive protection against tumorigenesis driven by CMV.

An intriguing disparity in overall health status between Whites, Blacks, and Hispanics and the CMV seroprevalence patterns by race and ethnicity in the U.S. evades a full explanation by factors of family size, household income level, education, marital status, census region, area of residence, country of birth or type of medical insurance. Differential exposure to CMV might partially explain these incongruities in health status, though [5,14,15].

The U.S. Hispano-Americans (Latinos) have significantly better health and mortality outcomes than the average population [16], contradicting their low socioeconomic status (SES). Medical experts have known for some time that Latinos living in the U.S. have on average a better life expectancy than non-Hispanic Whites. The so-called “Hispanic paradox” was recently supported by new data from the U.S. Centers for Disease Control and Prevention (CDC) [17-20]. Of note, the “paradox” seems not to hold true for follicular lymphoma (FL) and chronic lymphocytic leukemia (CLL) [21].

In this work we correlated age-adjusted incidence rates of all primary invasive cancers combined with the age-adjusted CMV seropositivity estimates among races and minorities in the U.S. The presentation of the reports used (**Table 1**) shows that, comparatively, CMV seropositivity is higher in the U.S. minorities than in non-Hispanic Whites. The significance of this observation is confirmed by statistical analysis. Moreover, we show that this effect is global, predominating across nations, societies, and histological types of malignant tumors (**Figs. 1-4** and **Table 3**).

**Table 1.**
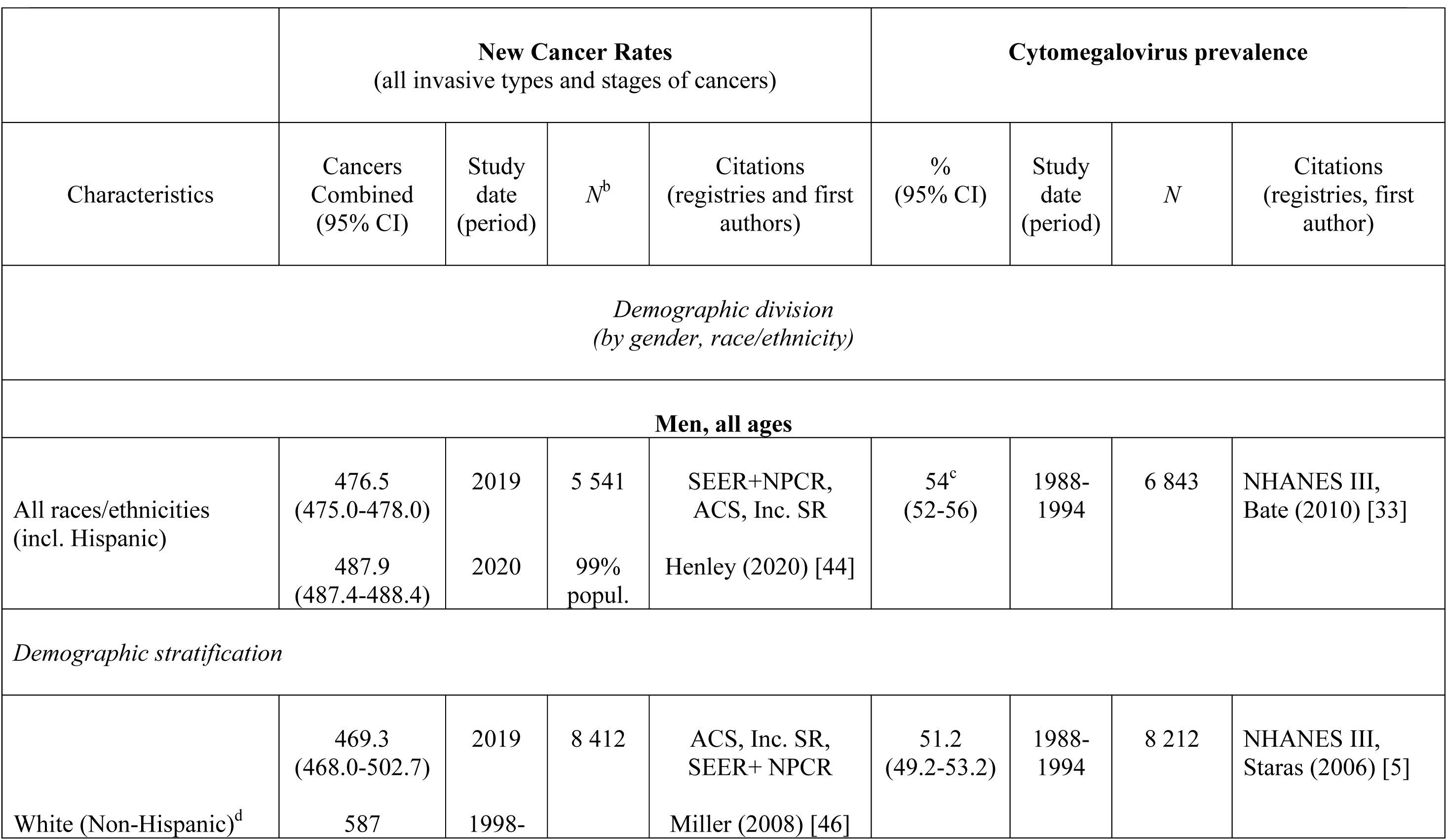
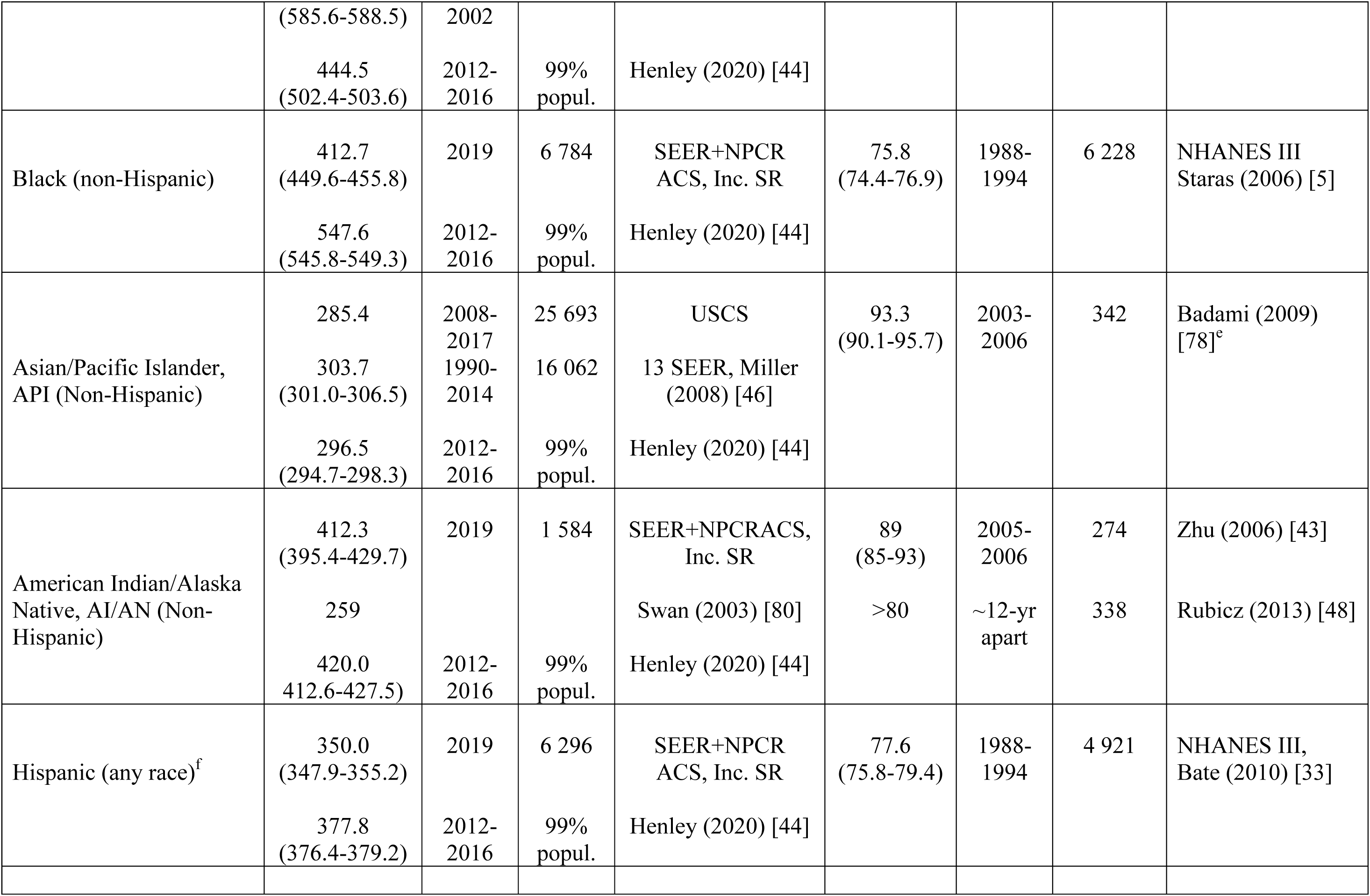
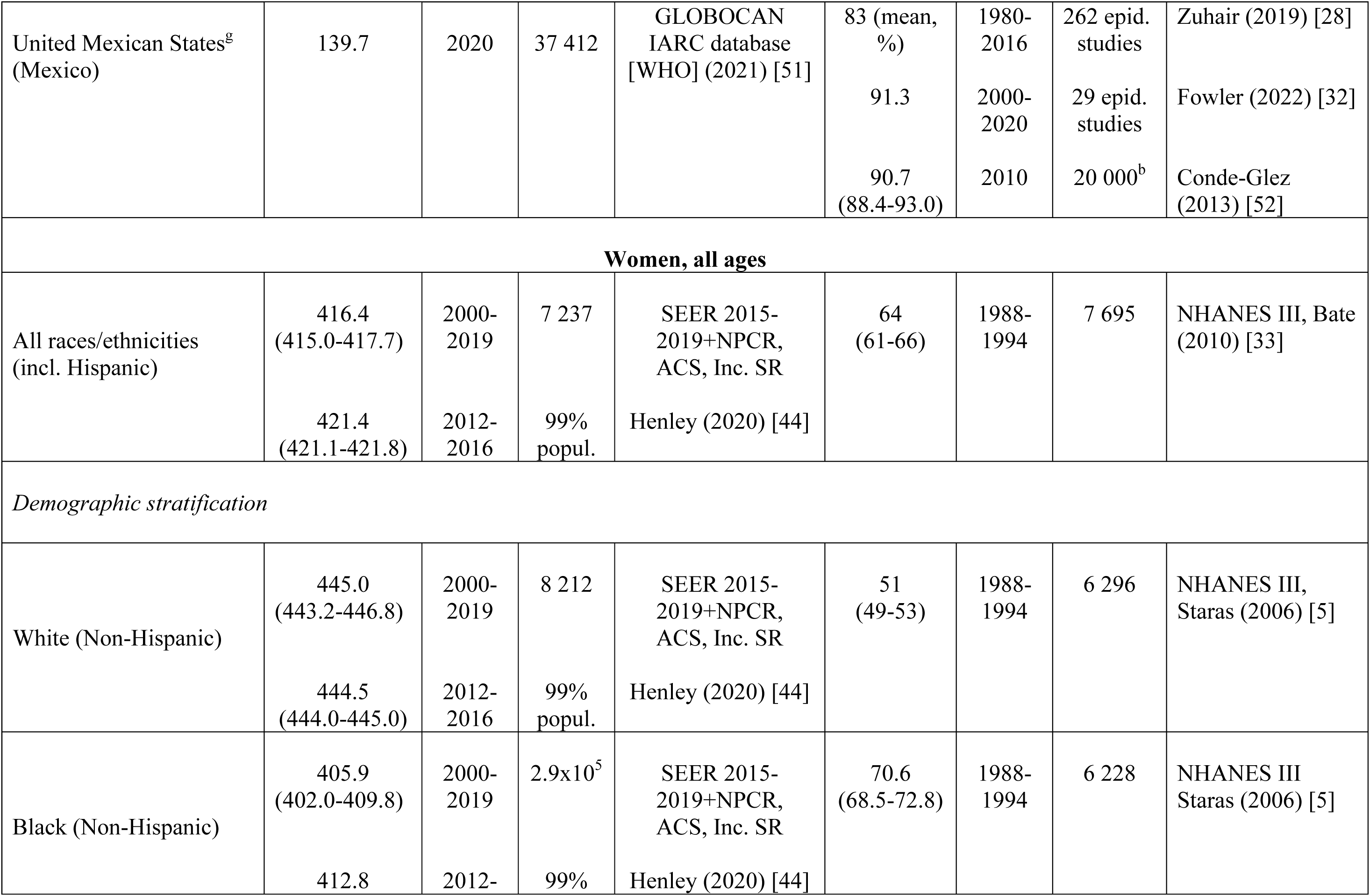
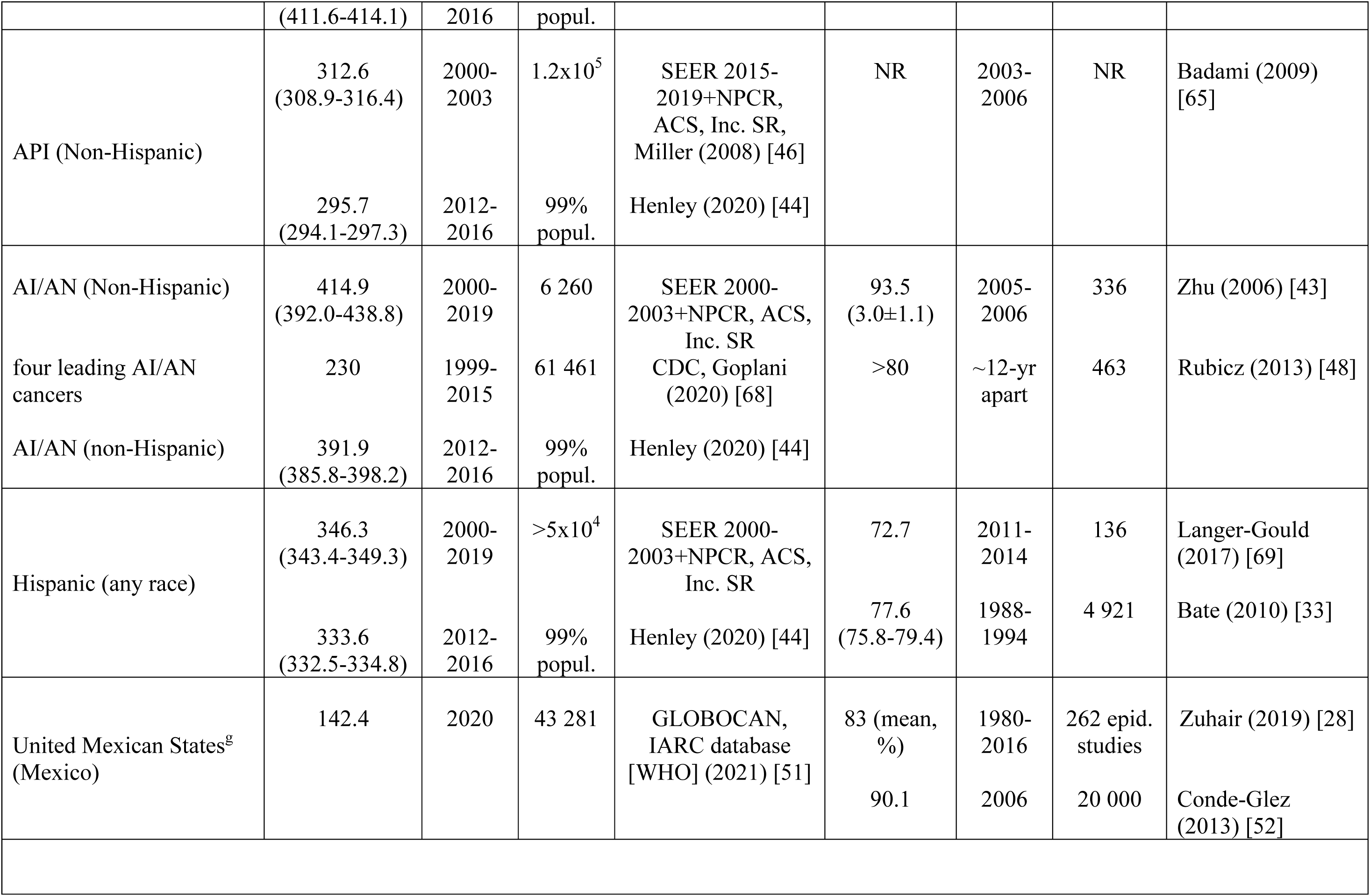
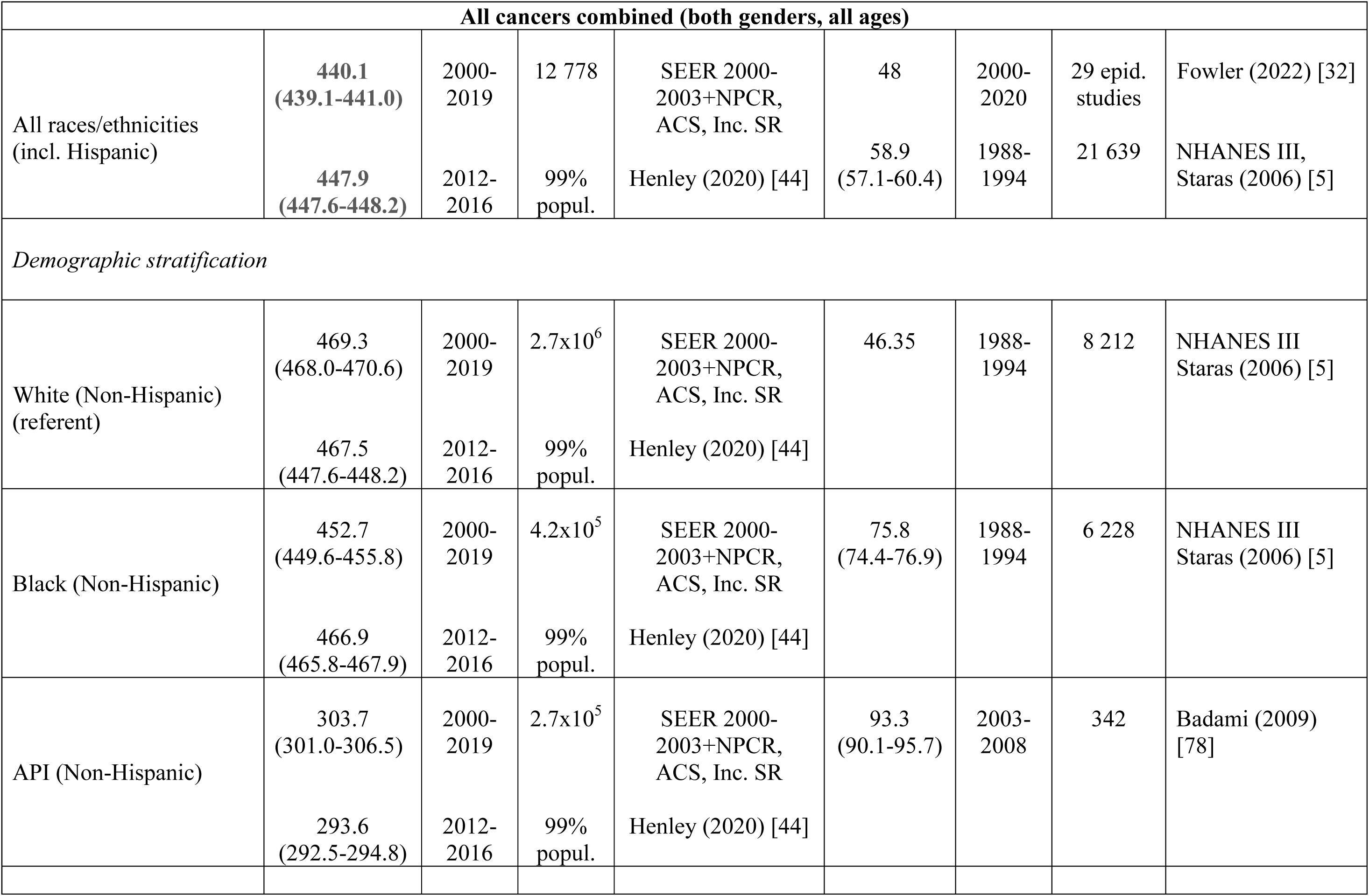
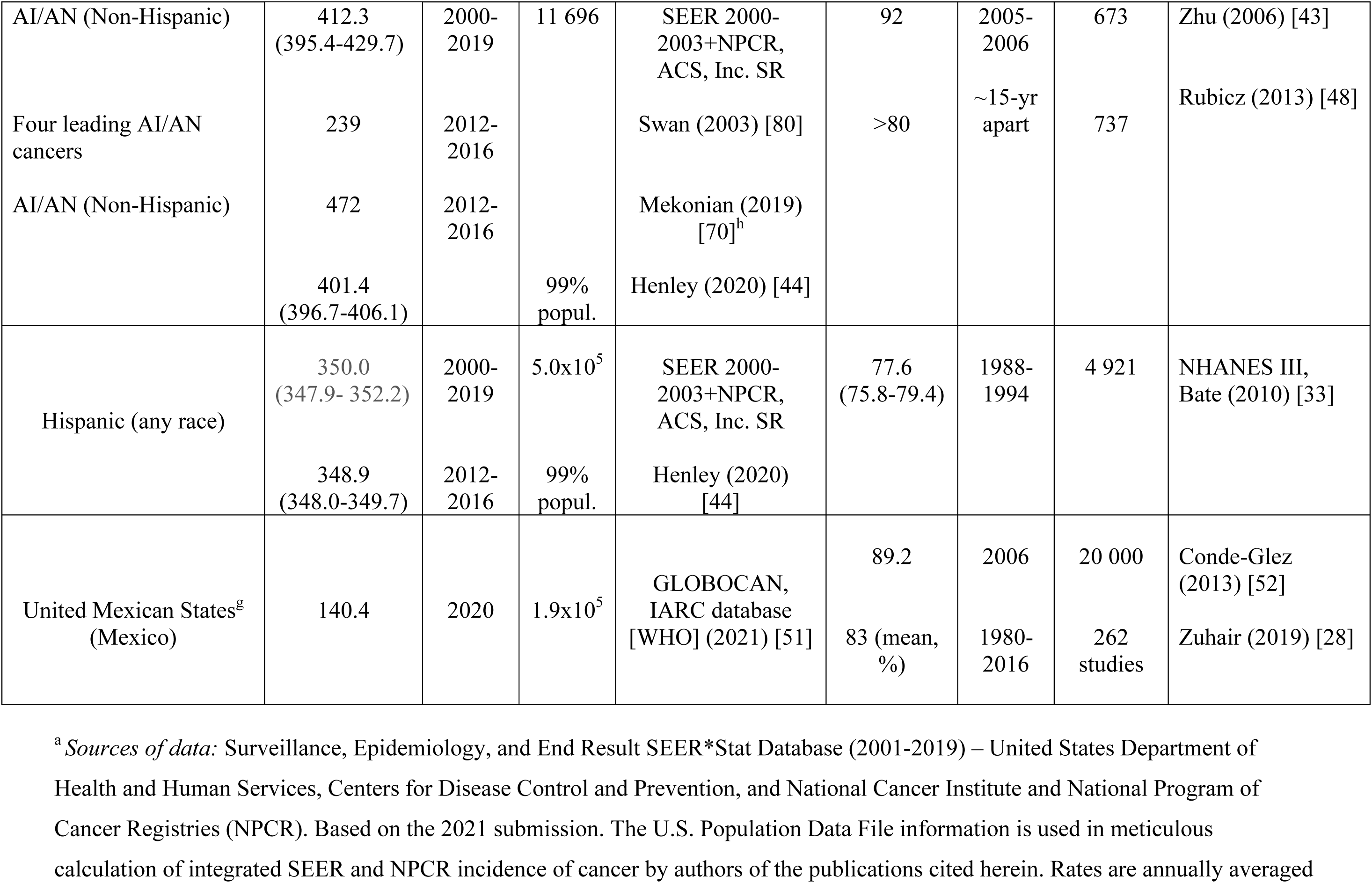
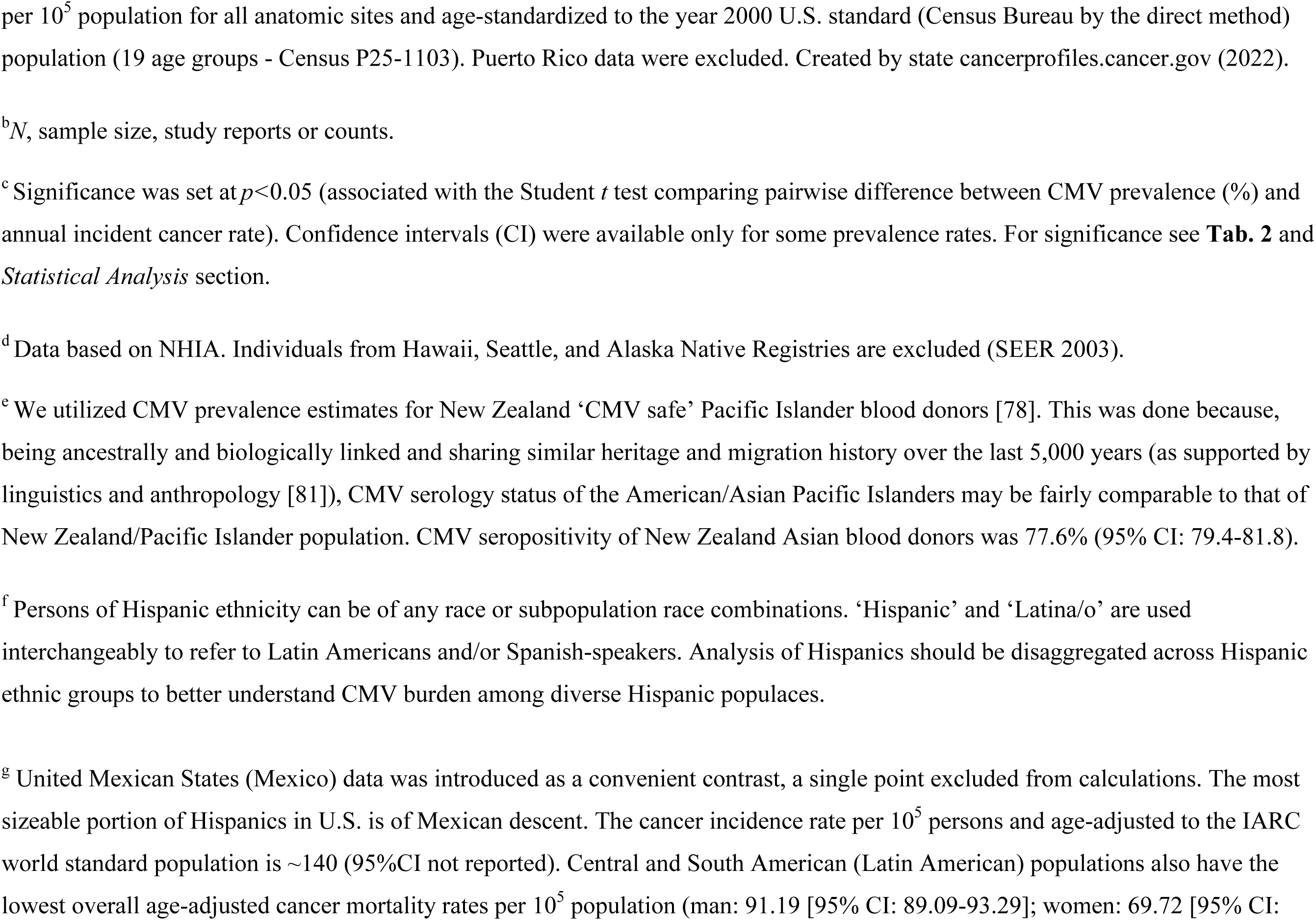
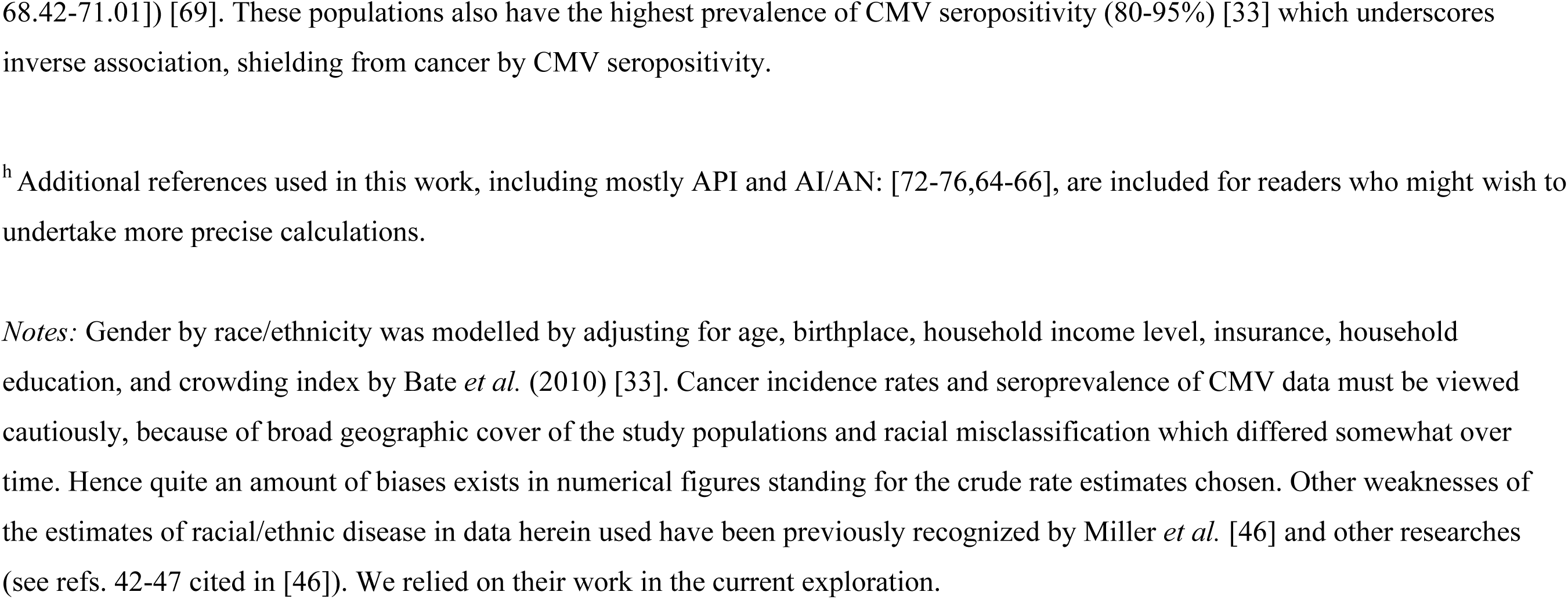
Race/ethnicity-specific epidemiology of new cancer cases [SEER 12 (2015-2019) + NPCR age-adjusted participants ^a^] and cytomegalovirus seroprevalence [NHANES III] (accessible data are selected for comparative purposes).

## Materials and Methods

### Study hypothesis

Our primary outcome of interest is an inverse association between rates of new cancer and CMV seropositive status the world over and within race/ethnic groups in the U.S.

### Study populations – worldwide data (73 countries)

In order to evaluate whether the proposed cancer incidence/CMV seroprevalence association holds true at a global scale, we accessed the World Health Organization Global Cancer Observatory (International Agency for Research on Cancer [IARC]), for data on worldwide cancer statistics. Worldwide annual incidence of new cancers (age-adjusted, both genders, all ages) was used. Country specific seroprevalence of CMV comes from work of Zuhair *et al.* [28].

### Study populations – the U.S

We examined a connection between an overall rate of new cancers (combined at all anatomical sites, all ages) among races/ethnic groups in the U.S. and the demographic pervasiveness of CMV seropositivity in these populations.

Patients with primary diagnosis of cancer (invasive) evidenced in the Surveillance, Epidemiology, and End Results (SEER) Program database between January 2007 and December 2015 were included in the study. The population was categorized into non-Hispanic Whites, non-Hispanic Black, Non-Hispanic Asian/Pacific Islanders (API), Non-Hispanic American Indian/Alaskan Natives (AI/AN), and Hispanics.

We collated national information from the SEER registry, an authoritative high-quality resource and error-proofed data source for the burden of cancer among the U.S. populaces. The SEER database is updated yearly. It contains patient demography, the primary site of the tumors, histology, and cancer stage at medical detection time [30,31]. The current study is based upon the reports of meticulous work of researchers on sure and accessible data on cancer rates for the U.S. race/ethnic groups (the Cancer Planet website and publications from the North American Association of Central Cancer Registries or NAACCR and the ACS). Oncologic parameters of race/ethnicity groups are defined by the American Joint Committee on Cancer (AJCC). Also, SEER program is the only source for historic population-based incidence and survival data (1975-2018). SEER 22 Incidence provided coarse rates (2000-2019) with the total registries data for all cancers combined, including sex, race, and ethnicity [34-50]. Numerical derivatives based on this data were made available by published reports herein referred to.

For data on CMV, we consulted a series of cross-sectional surveys drawn from NHANES collected by the National Center for Health Statistics (NCHS). The reports utilized here are based on the 2003-2004 wave which included CMV latency in the population and recognized consequences of persistent CMV infection on human health [29-31]. We made use of CMV prevalence data from the Third National Health and Nutrition Examination Survey (NHANES), 1988–1994. NHANES III was cross-sectional and stratified to allow for heterogeneity, multistage probability sample of civilian non-institutionalized U.S. population aged 2 months to 90 years.

To obtain current nationally representative estimates of the prevalence of CMV in the U.S., we used NHANES III study data (1988-1994) from Staras *et al.* [5]. NHANES is a series of cross-sectional surveys supervised and managed by the National Center for Health Statistics (NCHS) of the Centers for Disease Control and prevention CDC [38,39]. Also, we leaned on information and arguments from Bate *et al.* (**Tables 1, 2** and *Results* section [33]). The overall age-adjusted prevalence of CMV seropositivity seems not to have changed significantly in the U.S. for the intervals 1988-1994 and 1999-2004 (**Table 1** in [33]).

Also, we used query tools to collect literature related to CMV seroprevalence and cancer burden in the U.S. and elsewhere through MEDLINE, PubMed database search engine [terms ’cytomegalovirus’, ‘prevalence’, ‘IgG’, ’race/ethnic’, ‘global burden of cancer’, and their synonymous expressions]. Initially, we focused on CMV prevalence data for information regarding sex, race/ethnicity and SES, inversely concordant with a rate of cancer across geographic domains [13].

We have drawn on data from the primary literature, recognized reports and authoritative reviews on cancer incidence and CMV seroprevalence rates published heretofore (**Table 1**, [10]). The collection periods vary, spanning multiple years (see *Limitations*). We entirely relied on systematic reviews and meta-analyses of the epidemiological burden of CMV in the U.S. extracted from Medline and LILACS (Latin American and Caribbean Health Sciences Literature (10 October, 2020) [2,32,33].

### Statistical analysis

For the analysis of aggregated data points at the ethnicity level for CMV seropositivity and cancer incidence, we utilized available descriptive statistics to empirically determine the significance of correlation. For the CMV seropositivity variables (proportion *p* and the number of subjects *N*), we adopted the Bayesian framework with the assumed uniform U(0,1) prior and binomial Bi(p,N) data distribution yielding a beta Be(p*N+1, (1-*p*)*N*+1) posterior distribution. For the cancer incidence variables (estimated incidence and the 95% confidence interval (Henley *et al.* 2020 [44]) we assumed the maximum likelihood normal distribution; the Bayesian framework was omitted due to the variable methodology of incidence calculation and the assumption of a large number of observed data points. All the variables for all the ethnicity groups were independently simulated from their distributions 10 000 times under the null hypothesis that the data points are uncorrelated, and the Pearson coefficient of correlation was calculated each time. The empirical distribution of the correlation coefficient was estimated from the data giving the mean and 95% confidence interval. The 2-sided significance is calculated from the value of 0 and the significance level of 0.05. The analysis was done with custom scripts and the SciPy package of the Python programming language. Descriptive statistics, including frequencies and percentages were used for defining baseline characteristics of populations. Results are presented as counts, percentages (in parentheses) or median (interquartile 312 range) and frequency distributions depending on data type. Data was organized using Microsoft Excel software 2010 (Microsoft Corporation, Redmond, WA, U.S.). Spearman’s correlation coefficient (*ρ*) served to capture significance of correlation between variables assessed across the globe. Based on the method of covariance, it is a preferable method of measuring the agreement between variables of interest. Also, it provides information on the direction of the relationship. *P*<0.05 tested against an artifact of chance.

## Results

The prevalence of CMV seropositivity correlates inversely to cancer incidence rates across a broad range of histology and worldwide (**Table 3**; the top 3 malignancies with strongest correlations are represented in **Figs. 2-4**). Correlations are obtained in 30/34 (88.2%) tumors and are highly significant over 25/34 (73.5%) histology types. Also, an inverse association attains a significance if annual incidence of new cancers of all histologic types was combined (Spearman’s *ρ* = -0.732; *p<*0.001, **Fig. 1**). These results favor a possible oncoprotection that CMV infection provides to its host.

**Figure 2.**
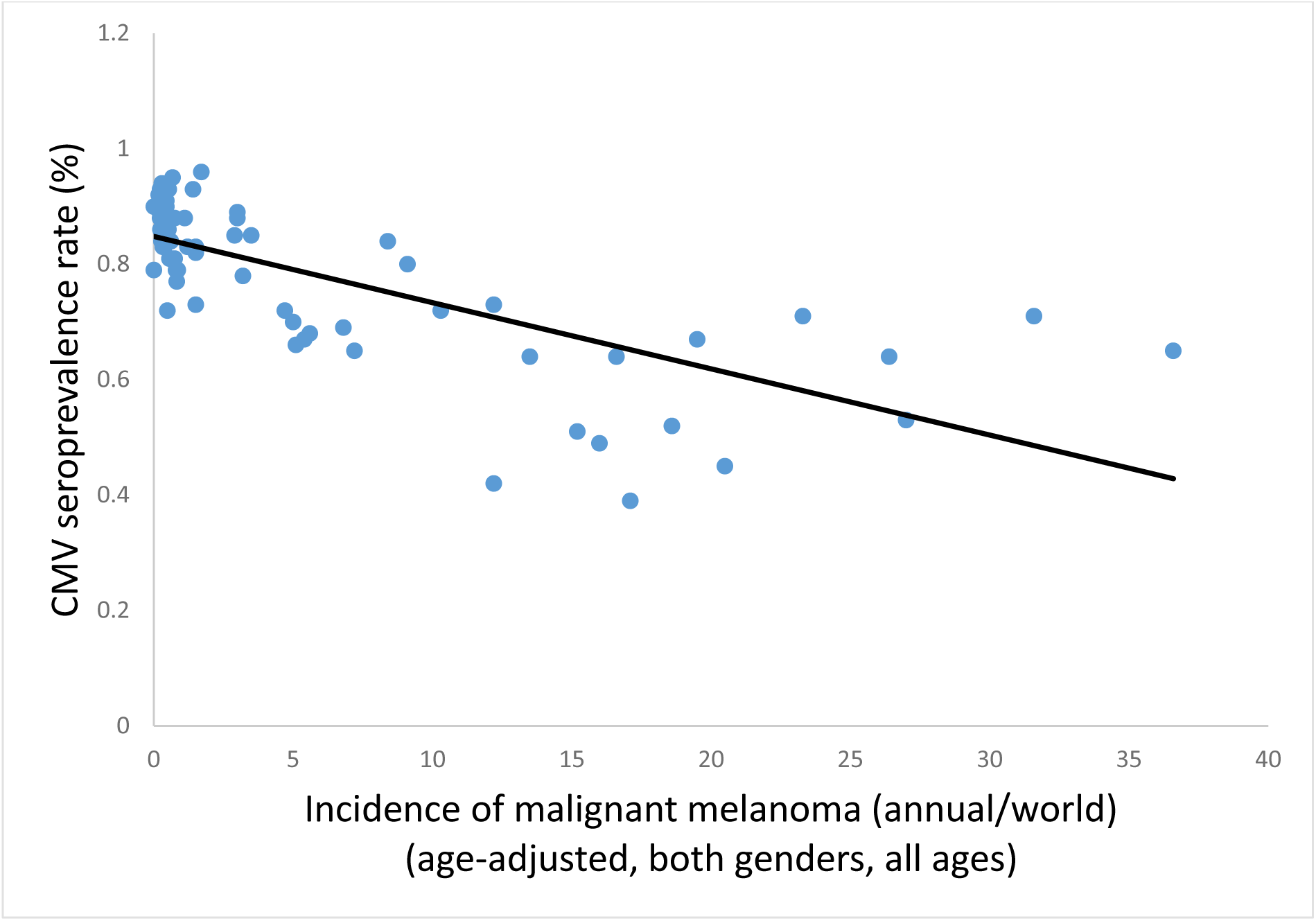
Malignant melanoma/10^5^ population *vs.* country specific prevalence of CMV seropositivity [28] correlate strongly inversely (Spearman’s *ρ* = -0.763; *p*<0.001) across 73 countries [51].

**Figure 3.**
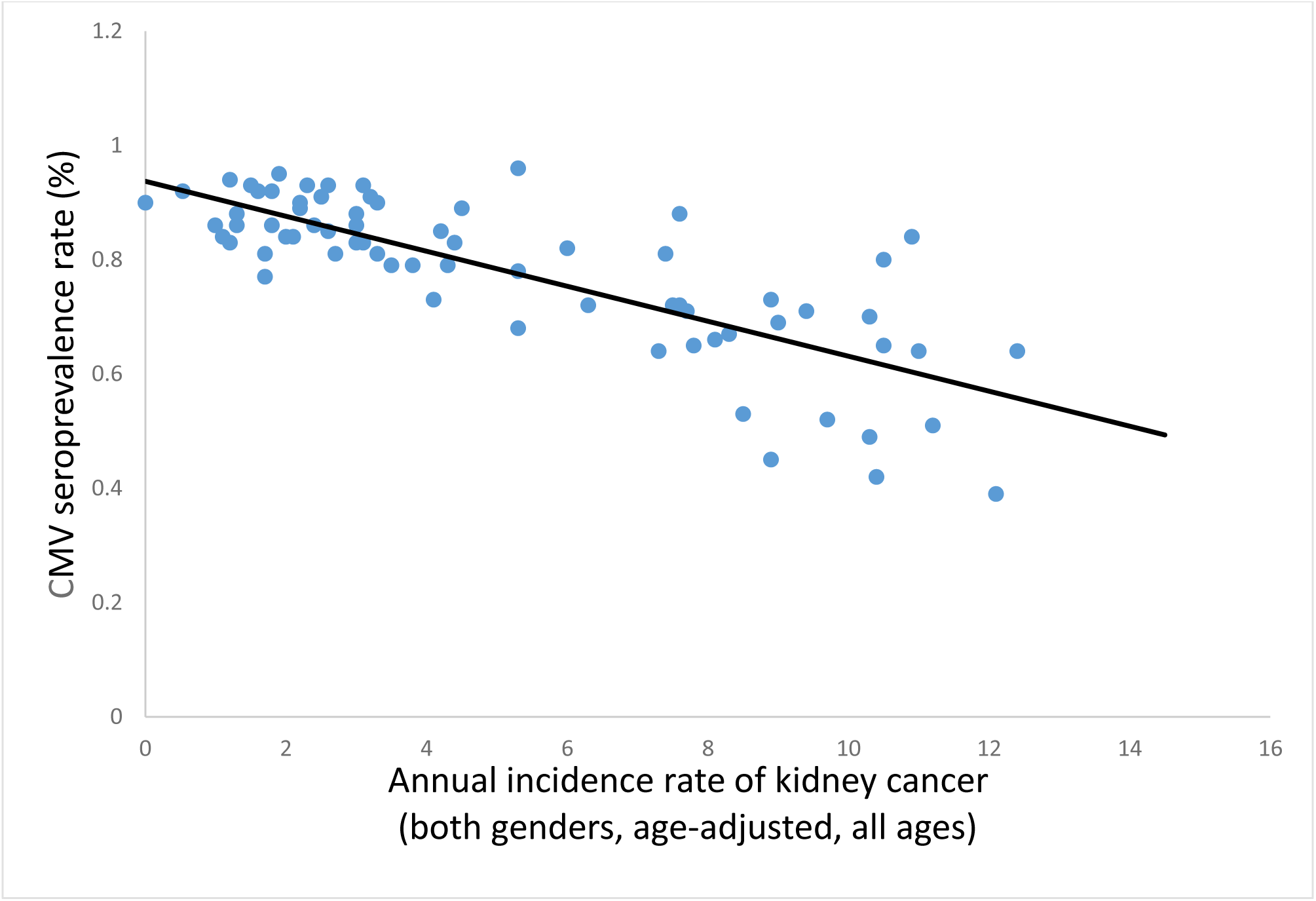
Kidney cancer/10^5^ population and the country specific CMV seroprevalence [28] are strongly and inversely connected across 73 countries [51] (Spearman’s *ρ* = -0.754; *p*<0.001).

**Figure 4.**
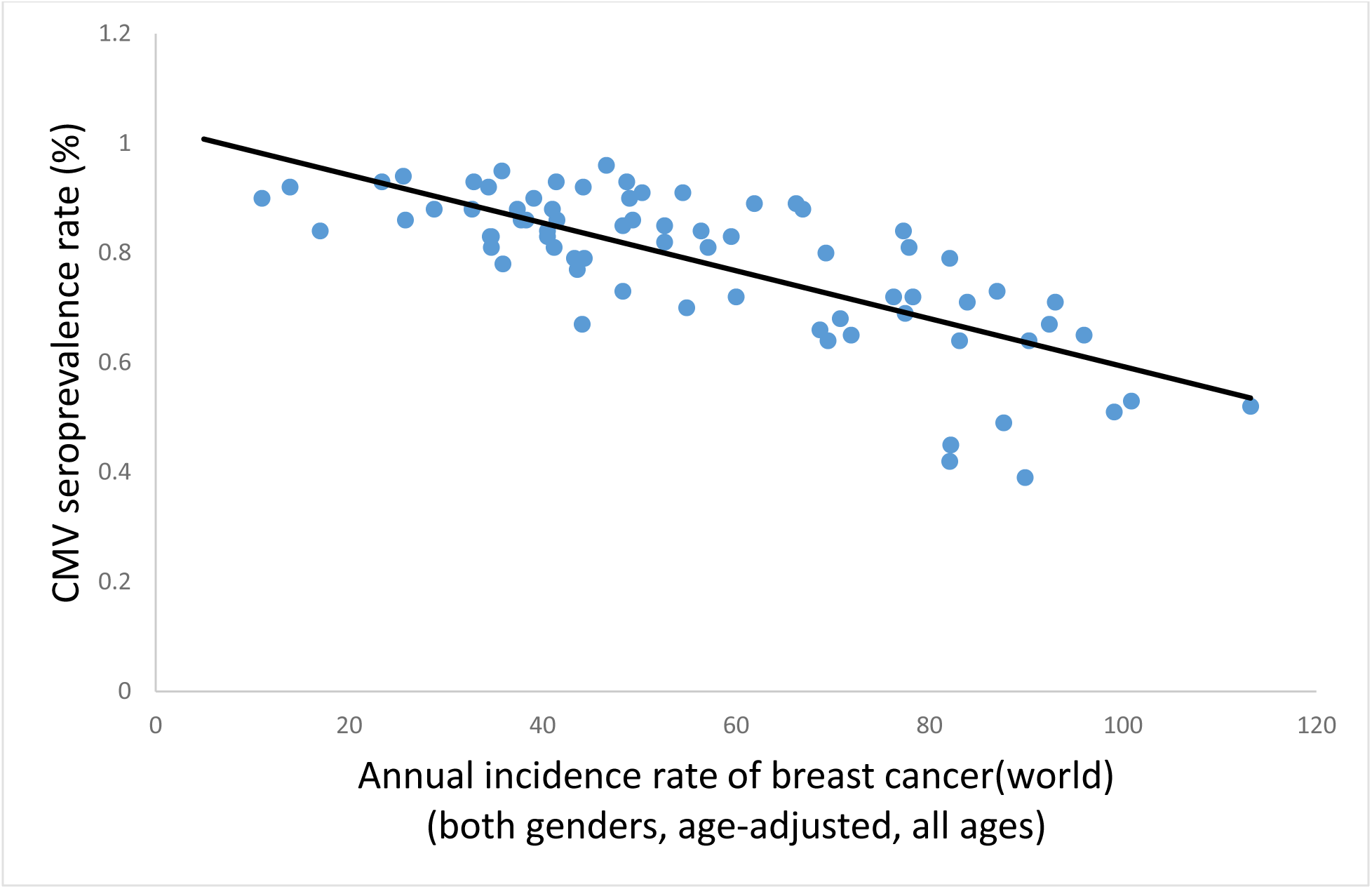
Incidence of breast cancer/10^5^ population and the country specific CMV seropositivity rate [28] spanning 73 continent-wide countries [51]. An inverse (protective) association is suggested by Spearman’s *ρ* = -0.719; *p*<0.001.

Conversely, CMV seroprevalence has a significant and co-incremental relation with an incidence of malignancies of nasopharynx and the gallbladder, suggesting a potential pro-oncogenic effect regarding these malignancies.

Notably, country-specific prevalence of CMV did not correlate inversely with an overall incidence of Kaposi’s sarcoma (**Table 3** and **Fig. 5**, Spearman’s *ρ* = -0.007; *p*=0.953). This is in line with a requirement of an operative T cell-mediated oncolytic response as being critical for the expression a protective capacity of CMV, supporting a hypothesis of specific T cell viral oncoprevention. We propose that a severely disrupted T cell immunity in HIV/AIDS patients, who comprise a vast majority of population with Kaposi’s sarcoma, effectively precludes a CMV-associated anti-tumor response.

**Figure 5.**
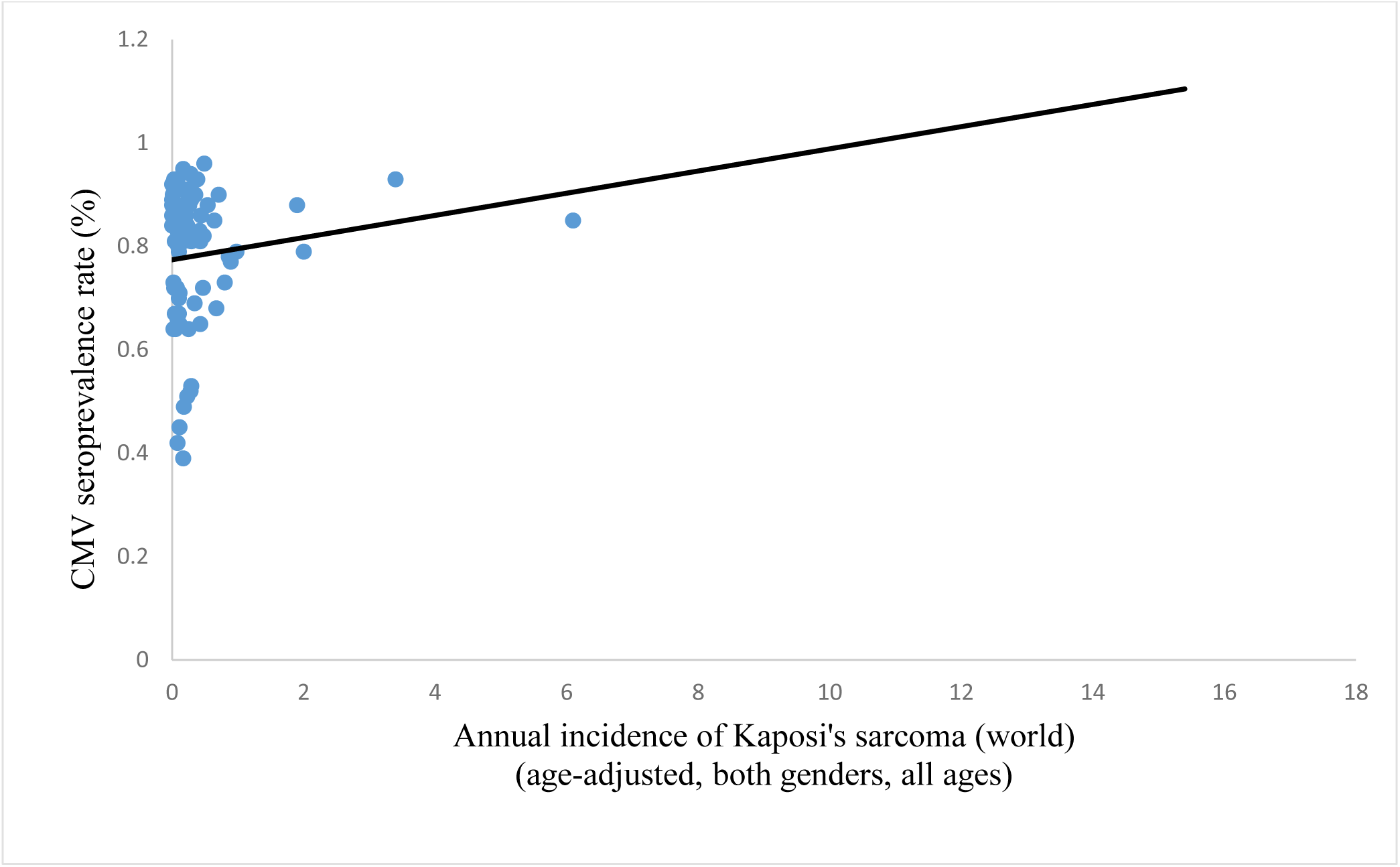
Incidence of Kaposi’s sarcoma (mostly in HIV+ people)/10^5^ population and the specific country-level rate of CMV seropositivity [28] covering 73 countries [51] are not correlated (Spearman’s *ρ* = -0.007; *p*=0.953).

**Table 1** exhibits comparative data on race/ethnic estimates of the incidence of all cancers combined (both genders, all anatomical sites, all ages) in the U.S, and the territorially matching prevalence of CMV seropositivity. The estimate of cancer incidence for Mexico is included as a contrasting population but was discarded from statistical calculations. The aggregated results of studies mentioned therein provide a clear representation of a higher CMV pervasiveness in the U.S. minorities than in non-Hispanic Whites.

**Table 2** represents a statistical analysis of the SEER data on racial/ethnic rates of cancer (Henley *et al.* [44]) and the corresponding prevalence of CMV. Also pointed in **Table 2** is a highly significant and inverse correlation (*p*<0.001; mean: -0.674; SD=0.018; CI=-0.711) between the standardized cancer incidence and the CMV seroprevalence in the U.S. An outlying group are the Eskimos (AI/AN), perhaps because of an impaired T cell function which distinguishes this ethnicity from the others [66,67]. Consequently, CMV-specific cytotoxic T cell oncolysis in Eskimos is not an efficient suppressor of clonogenic processes (see *Discussion*). The results also support broad evidence (Figs. 1-4) suggesting a tendency of CMV seropositivity to monotonically decrease as SES improves across a range of developed and underdeveloped countries. This may explain a higher incidence of malignancies in economically progressive countries.

**Table 2.**
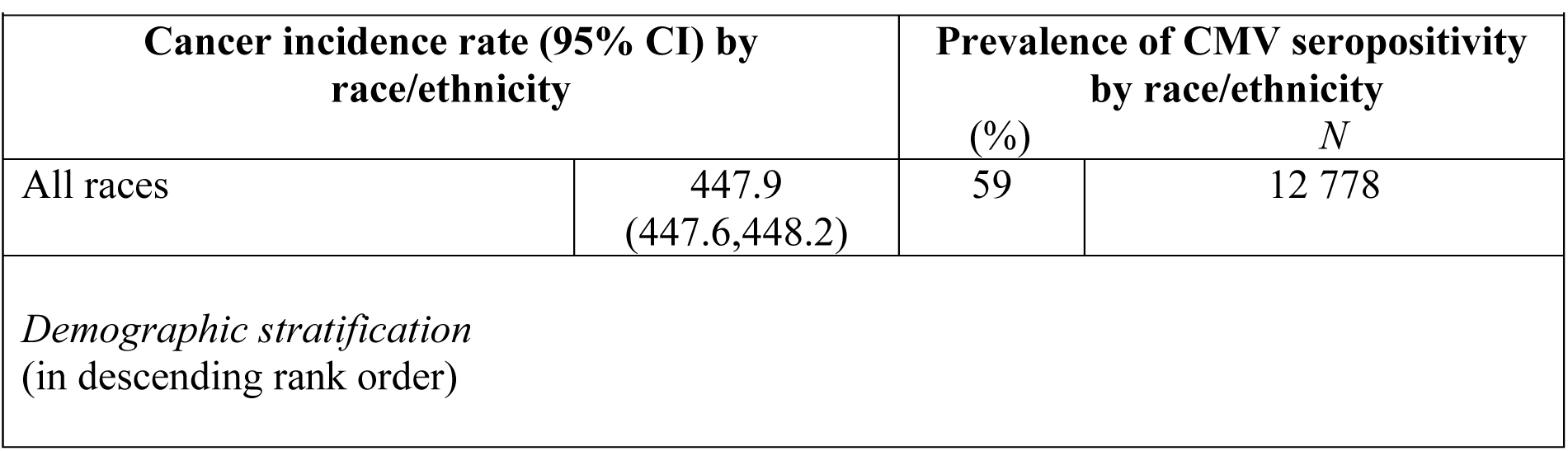
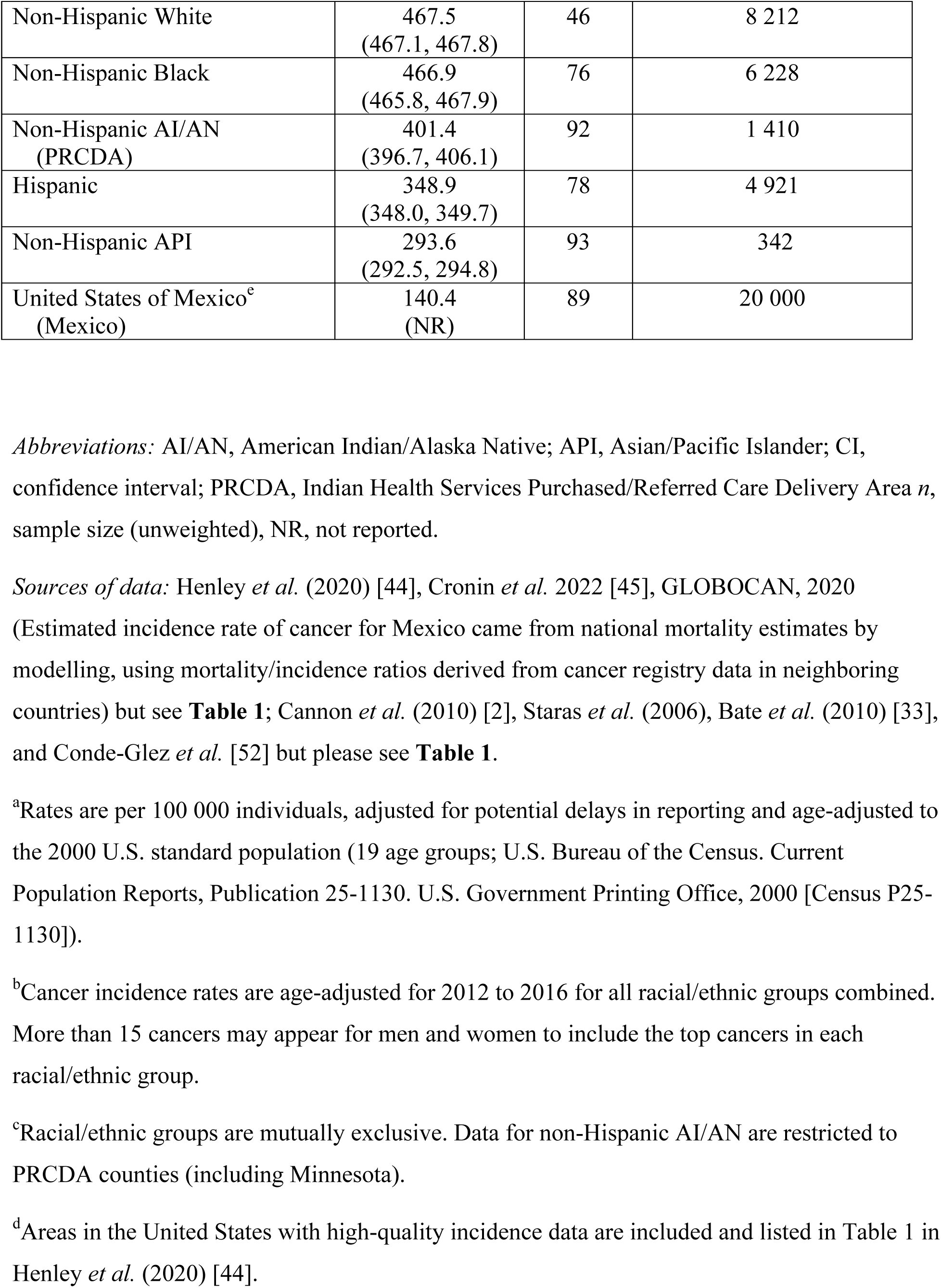
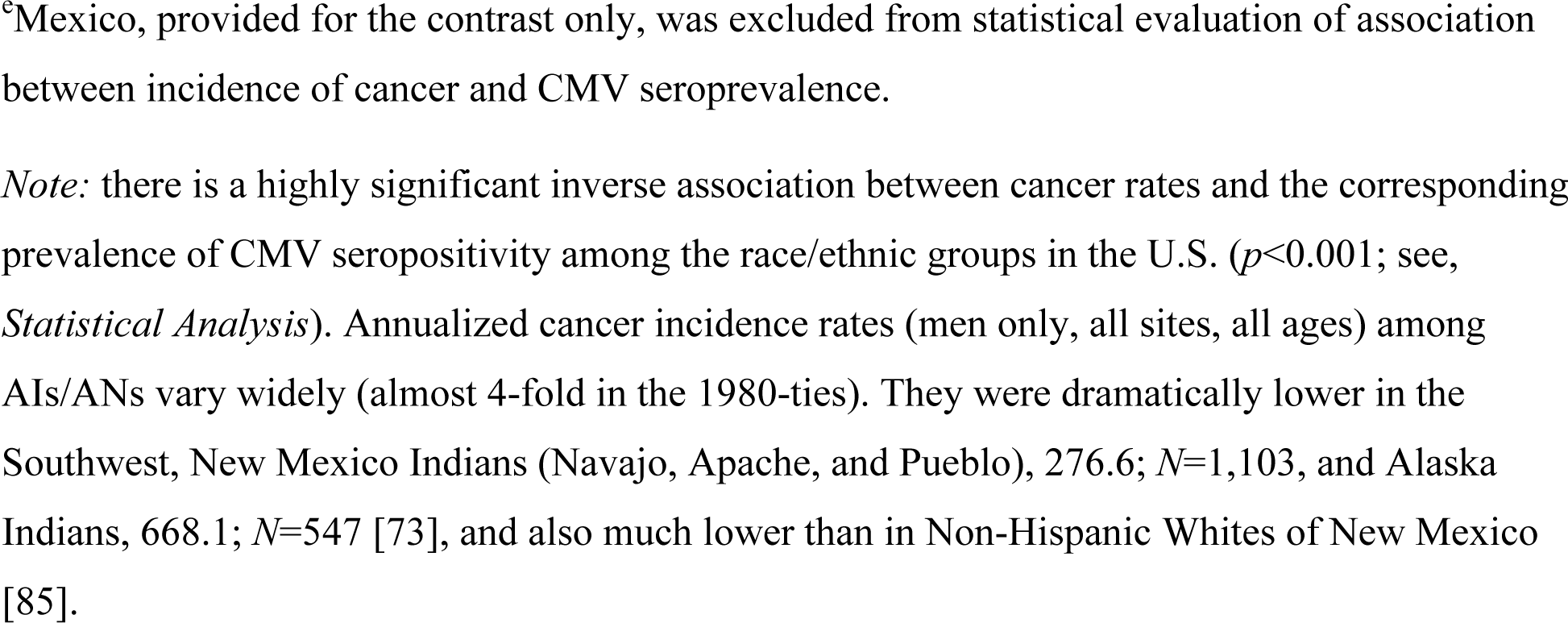
Age-standardized, delay-adjusted incidence rates ^a^ for the most common cancers ^b^ (all anatomical sites combined, both genders, all ages) among the racial/ethnic groups ^c^ (2012-2016)^d^ in the United States compared to their CMV seropositivity (%).

## Discussion

Cytomegalovirus is wreathed in mystery, a Gordian knot of biological knowledge and understanding [82]. Cytomegalovirus had evolved a peculiar virus-host symbiosis by a taciturn tactic that co-opts or escapes immune pathways in order to arrange its persistence in the host. A strong inverse statistical correlation between the incidence of cancer and CMV seroprevalence covers quite a swath of cancer spectra and territorial ranges. Although correlation does not categorically infer causation, this one may underscore a causal meaning. Besides the benefits of vaccine against CMV [53], it may suggest an impeded carcinogenesis by generating the CMV-induced tumoricidal T cells. CMV-based vaccine antagonistic to tumorigenesis would possibly result in a global regression of cancer. Comparing non-Whites to Whites, Cannon *et al.* [2] evaluated that CMV seroprevalence is consistently 20-30 percent points higher in the former than in the latter (summary PR=1.59, 95% CI=1.57-1.61). Some ethnic groups had CMV seroprevalence ∼ 100% (Fig. 4A in [2]). In agreement with the current work, Fowler *et al.* [32] discounted the importance of ethnicity *per se* as a risk factor for the CMV infection. Similar conclusions were reached by Lantos *et al.* [54], Rook [55], and Dowd *et al.* [56,57].

Elimination of health disparities as a consequence of racial and ethnic differentials has been earlier recognized as important [47,48,83]. Socioeconomic disparities across race/ethnicity categories impact the level of CMV seroprevalence [83]. These may have biased lower estimates of cancer incidence at the time. Rather than the race/ethnic divides of themselves, we propose CMV infection as an oncopreventer both of some hematologic malignancies at our clinic and of cancers worldwide [13]. Although CMV prevalence and education level, SES, and household income are associated with race and ethnicity, we speculate here that latent CMV infection is a fundamental cause underlying disparities in cancer incidence among race/ethnic groups in the U.S. and worldwide. The NHANES III data (the U.S. 2011-2012) report CMV prevalence (race/minority) in children 1-5 years of age as 37% among non-Hispanic other/multiracial, 31% among Hispanic, 15.9% among non-Hispanic Black, and 10.6% among non-Hispanic White ethnicities [33]. This data is inversely proportional to the incidence of all cancers (combined) in populations of these youths, indicative of a higher risk of cancer in CMV seronegative populations in the U.S. Also, unlike Hispanos, cancer rates in Cubans were comparable to non-Hispanic Whites, and Puerto Ricans and Cubans in Florida had rates of some solid cancers similar to non-Hispanic Whites despite the rates of these cancers being significantly lower in their countries of origin [22]. High incidence of cancer despite a high prevalence of CMV in the Inuit (Eskimos) may be a consequence of deficient T cell immunity in this ethnic population [23,24]. This is to be expected and, indeed, we found no global correlation between the incidence of Kaposi’s sarcoma (mostly diagnosed in HIV-positives with compromised T cell immunity) and the prevalence of CMV (**Fig. 5**). CMV does not exert protection unless T cell immunity is functional.

Immigrants to U.S. experience decreasing incidence rates of cancer of infectious origin (hepatitis B virus, *Helicobacter pylori*, human papillomavirus) which are prevalent in their countries of origin. On the contrary, incidence rates of lung, breast, colorectal and prostate cancer have been on the rise despite remaining relatively low in the host nations [57-59].

The statistical analysis indicates an inverse correlation between CMV pervasiveness and the race-specific cancer incidence, a valuable hint at a possible oncoprotective effect of the pathogen. Previously, we speculated that CMV may confer a protection against B cell dyscrasias [13]. For example, there is a highly significant inverse link (**Fig. 1**. Spearman’s *ρ* = -0.754; *p*<0.001) between all invasive cancers combined (both genders, all ages) and the country specific CMV prevalence profile across the mainland and sea-coastal regions of 73 countries (**Table 3**). We draw attention to a possible protective effect of the CMV infection as an unappreciated factor which may subvert oncogenesis.

**Table 3.** Correlation between worldwide incidence rate (100,000 population/year) of individual malignant tumors across 73 countries [WHO’s GLOBOCAN database (51)] and the country specific prevalence of CMV seropositivity rates (28).

| Tumor/localization | Spearman's<br>$\rho$ | $p$-value | Inverse<br>correlation (+/--) |
| --- | --- | --- | --- |
| Melanoma (skin) | -0.763 | 0.001 | + |
| Kidney | -0.754 | 0.001 | + |
| All cancers | -0.732 | 0.001 | + |
| All cancers (excluding non-melanoma skin cancer) | -0.726 | 0.001 | + |
| Breast | -0.719 | 0.001 | + |
| Testis | -0.711 | 0.001 | + |
| Non-melanoma (skin) | -0.692 | 0.001 | + |
| Colorectum | -0.671 | 0.001 | + |
| Vulva | -0.665 | 0.001 | + |
| Prostate | -0.663 | 0.001 | + |
| Corpus uteri | -0.656 | 0.001 | + |
| Oropharynx | -0.651 | 0.001 | + |
| Pancreas | -0.633 | 0.001 | + |
| Multiple myeloma | -0.633 | 0.001 | + |
| Leukemia | -0.632 | 0.001 | + |
| Hodgkin lymphoma | -0.618 | 0.001 | + |
| Non-Hodgkin lymphoma | -0.617 | 0.001 | + |
| Mesothelioma | -0.574 | 0.001 | + |
| Lip, oral cavity | -0.551 | 0.001 | + |
| Lung | -0.548 | 0.001 | + |
| Brain/CNS | -0.541 | 0.001 | + |
| Thyroid | -0.532 | 0.001 | + |
| Bladder | -0.519 | 0.001 | + |
| Ovary | -0.461 | 0.001 | + |
| Penis | -0.432 | 0.001 | + |
| Hypopharynx | -0.377 | 0.001 | + |
| Salivary glands | -0.35 | 0.002 | + |
| Gallbladder | 0.316 | 0.006 | -- |
| Nasopharynx | 0.266 | 0.023 | -- |
| Vagina | -0.224 | 0.056 | + |
| Larynx | -0.165 | 0.164 | + |
| Esophagus | -0.149 | 0.208 | + |
| Cervix uteri | 0.118 | 0.319 | -- |
| Stomach | -0.085 | 0.473 | + |
| Kaposi's sarcoma | -0.007 | 0.953 | + |
| Liver | 0.007 | 0.951 | -- |

*Notes:* Significant inverse correlation, implying a possible prevention of tumorigenesis due to primary CMV infection the world over, is marked by +; the apparent lack of this association (i.e. absence of CMV-mediated anti-cancer protection) is denoted by --. Most recent clinical studies confirm negative (i.e., protective) correlation between CMV infection and both colorectal [86] and bronchogenic cancer [87].

We sought to grasp a better understanding of fluctuating incidence of new cancers among immigrant and established indigenous populations by consulting relevant reports. We envision that dilution of the prevalence of CMV, due to a progress in economic opportunity and an open access to competent medical patronage, may have resulted in increased cancer rates, indeed the epidemics of neoplasms [60]. Societal development aside, poverty remains a considerable medical concern [30,33,62,72]. Higher prostate and lung cancer rates are reported in established immigrant enclaves from Japan and China in the U.S. than are found in Japan and China, probably because improved SES in the U.S. (i.e. a better hygiene and improved medical care) with a consequent decline of CMV-mediated oncoprotection in the U.S. Also, individuals with low SES have higher antibody titers to CMV [31] and presumably more protection against cancer.

Crucially, CMV induces a specific, CMV-determined, T cell mediated antitumor effect in immunocompetent persons but fails in patients with inoperative T cell immune surveillance, like Kaposi sarcoma (**Fig. 5**). The role of CMV acquisition and the consequent T lymphocyte-specified inhibition of tumorigenesis may prove of importance in pre-empting various malignancies, particularly those that can be detected at an early stage, such as breast and colon cancers.

Disaggregation (decomposition) of ethnic data in racial/multiethnic studies of cancer *vs.* rates of cancer cases combined and the CMV prevalence may, while quite information-rich, also be a source of bias due to unmeasured confounders or mediators [79]. Disaggregation may mask true biological linkages which would be more accented if global bulk of data were analyzed. Hoshiba *et al.* [61] examined the long-term (1980-1998) dynamics of CMV seroprevalence in pregnant women in Japan. Complement-fixing antibody and specific IgG antibody, as measured in sera, decreased gradually from 93.2% to 66.7%. CMV-IgG seropositive rates were 87.4% in 1985, and 75.2% in 1996 to 1997. This provides a separate hint at a possible protective effect of CMV in this population. Of note, the incidence of cancer increased in parallel to decrease of CMV seropositivity in this Japanese population.

Most recent clinical research on colorectal [86] and bronchogenic cancer [87] confirm our evidence for the protection against cancer (**Table 3**) conferred by the CMV infection.

## Limitations

Notable limitations of this work are difficult to lift. A major limitation are the data used from literature and reviews with varying collection periods which span numerous years (**Table 1**). Disparate time scales occasionally underlie epidemiological studies of CMV in the low-income greater neighborhoods. There, the prevailing cultural settings and areas of higher deprivation (with highest prevalence of CMV infection) frequently overlap confounding the measured estimates (**Table 2**). Small numbers yield unstable prevalence rates. The roundabouts of more disadvantageous neighborhoods, culturally and genetically distinct yet sharing remotely entangled backgrounds, often overlay and may affect accuracy and consistency of the estimates. Race and ethnicity were differently categorized by the investigators. Also, the options were defined by study participants. Marked disparity between African Americans and White patients might have resulted from different indication settings for CMV testing and in unknown proportions. Also, there still remains a possibility that, besides the CMV, there could have existed yet another causative tumor-suppressive agent. Consequently, formal adjudication of causality of statistical significance is not allowed by the cross-sectional nature of the data in many studies we used. Still, prospective studies suggest that causality could be at work behind significance of statistical tests; high degree of disparity between Blacks and Whites is unlikely to result solely as an artifact of testing biases. Pronounced variance in cancer incidence in AI/AN reported by Henley *et al.* [44] and Cronin *et al*. [45] are possibly due to improved financial attainment and hygiene, cleaning, and the privilege of preventive medical care although a drop of CMV prevalence in this populace has not yet been accurately registered. The New Zealand Islanders’ CMV seropositivity estimates presented here (**Table 1**) may crudely parallel those in APIs who live on the U.S. Pacific Island Territories rather than that of API migrants living on the U.S. mainland. The estimates should be considered with circumspection and checked with regard to future vaccination strategies to fight CMV infection [64]. We have to take the results presented here within the context of these limitations.

## Conclusion

Our work is the first to analyze demography of cancer incidence rates relative to the prevalence of CMV seropositivity both in U.S. race/ethnic groups and worldwide. A biological connection between the ubiquity of CMV and the incidence of neoplasms is predicated by a worldwide correlation between the two. This result may further inspire an oncologic initiative for development of the CMV-based antitumor vaccine constructs that could result in a significant reduction of human cancers the world over. Here, we aimed at just this.

## Data Availability

All data produced in the present study are available upon reasonable request to the authors

## Acknowledgements

We credit the staff of the SEER and NHANES III who disseminated publicly visible data, essential for our analysis, and acknowledge the funding by the Ministry of Science, Technological Development and Innovation (MSTDI) of the Republic of Serbia. The funder did not play a role in the design of the study; the collection, analysis, and interpretation of the data; the writing of the manuscript; and the decision to submit the manuscript for publication.

## Authors’ contributions

Conceptualization: MJ, AK, TJ, MT-B.

Funding acquisition: TJ, BM, MT-B.

Writing — original draft preparation: MJ, AK, IĐ, OM.

Writing — review and editing: MJ, TJ, IÐ, OM, BM.

Investigation and Methodology: MJ, OM, AK.

Project administration, Supervision, Resources: TJ, AK, BM, and MT-B.

All authors read, critically revised, and approved the submission of the final version of the manuscript.

## Data availability statement

All Data are available from the corresponding author upon reasonable request.

## DECLARATIONS

### Ethics approval and consent to participate

Not applicable.

